# Predicting Respiratory Decompensation in Mechanically Ventilated Adult ICU Patients

**DOI:** 10.1101/2022.02.01.22270119

**Authors:** Yvette Tan, Michael Young, Akanksha Girish, Beini Hu, Zina Kurian, Joseph L Greenstein, Han Kim, Raimond Winslow, James Fackler, Jules Bergmann

**Author notes:** These authors contributed equally (Yvette Tan, Michael Young).

## Abstract

**Background:** Mechanical ventilation is a life-saving treatment in the Intensive Care Unit (ICU), but often causes patients to be at risk of further respiratory complication. We created a statistical model utilizing electronic health record and physiologic vitals data to predict the Center for Disease Control and Prevention (CDC) defined Ventilator Associated Complications (VACs).

**Methods:** We constructed a random forest model to predict occurrence of VACs using health records and chart events from adult patients in the Medical Information Mart for Intensive Care III (MIMIC-III) database. We trained the machine learning models on two patient populations of 1921 and 464 based on low and high frequency data availability. Model features were generated using both basic statistical summaries and tsfresh, an automated python feature generation library. Classification to determine whether a patient will experience VAC one hour after 36 hours of ventilation was performed using a random forest classifier. Two different sample spaces conditioned on five varying feature extraction techniques were evaluated to identify the most optimal selection of features resulting in the best VAC discrimination. Each dataset was assessed using K-folds cross-validation (k = 10), giving average area under the receiver operating curves (AUROC) s and accuracies.

**Findings:** After feature selection, hyperparameter tuning, and feature extraction, the best performing model used automatically generated features on high frequency data and achieved an average Area Under Receiver Operating Characteristic Curve (AUC) of 0·83 ± 0·11 and an average accuracy of 0·69 ± 0·10.

**Interpretation:** We constructed a promising model to predict VACs 1 hour prior to occurrence with 36 hours of ICU patient data. The model provides early warnings of VACs, which may allow actionable therapies to prevent or mitigate ventilator associated complications.

**Funding:** N/A

## INTRODUCTION

Mechanical ventilation is a life-saving treatment for patients with respiratory failure. Every year in the U.S., up to 800,000 patients receive mechanical ventilation treatment (1). However, ventilated patients are at risk of further respiratory complication that increases the longer a patient is mechanically ventilated (2). A major obstacle in identifying vulnerable patients is the lack of an accurate and objective clinical definition for respiratory decompensation in the context of mechanical ventilation. In 2013, Magill et al introduced a framework for identifying ventilator-associated events (VAE) in adults (2). This VAE framework (3) is a surveillance definition based on objective ventilator settings and measures used to monitor the incidence of pulmonary incidents caused by medical treatment in ventilated patients (4). The VAE framework has three layers; the first specifies ventilator-associated complications (VAC), and the second and third layers further label patients with infection-related ventilator-associated complications (IVAC) and presumed ventilator-associated pneumonia (PVAP) respectively (3). The VAE framework’s diagnostic criteria for a VAC is used as a proxy definition for respiratory decompensation in mechanically ventilated patients (1). This definition enabled us to have a primary endpoint for our classifier.

Standard clinical practice dictates that patients are maintained on the lowest possible ventilator settings to provide adequate oxygenation and patients are gradually weaned from ventilation as soon as possible. A complication is typically detected when ventilator settings are increased for a significant length of time. A VAC is an event of pulmonary deterioration that a patient may experience while receiving mechanical ventilation treatment (4). VACs can be caused by acute lung injury, pneumonia, pulmonary edema, and acute respiratory distress syndrome (ARDS). The occurrence of a VAC necessitates sustained reliance on ventilator support, which often increases ICU length of stay and exacerbates both the already steep costs of hospital stays and a patient’s mortality rate and risk of comorbid complications (5)

A clinically significant early warning of decompensation could trigger timely investigations that either decrease risk or its potential to harm the patient. Techniques used for critical care assessment such as the VAP (ventilator associated pneumonia) definition are insufficient to predict mechanical ventilation-related events before a physician detects obvious physiological signs of a complication due to factors such as variability in progression of patient conditions and difficulty in application of definitions to patient cases (6). In this study, we present a predictive, early-warning classifier model for ventilator-associated complications as a proxy definition for respiratory decompensation.

### Aims

We built predictive models to assess risk of VAC using patient demographics, EHR data, and physiologic time series data.

Our study aims were two folds:

1. Build a model to predict VACs one hour before occurrence using 35 hours of ventilation data
2. Evaluate different feature extraction techniques on our model

In this study, we wanted to compare different feature extraction techniques on different granularities of data. In particular, we looked at manual statistical features vs. automated feature extraction, and high-resolution vs low-resolution data.

In order to create our models, we leveraged data from MIMIC-III, an open-access database (7), and utilized machine learning techniques to construct various models. Our results have the potential to inform clinicians on the need for timely critical interventions during the ICU stay of mechanically ventilated patients before obvious signs of decompensation. With a clinically significant early warning system, the monitoring physician will have a risk score, in addition to other physiological measurements, as evidence to make changes to treatment protocol if necessary. Earlier intervention on the part of the clinician can help mitigate and altogether prevent complications that have been predicted to occur. The prevention of a VAC improves patient outcomes by shortening duration of ICU stay, reducing hospital costs, and preventing further mechanical injury (8).

### Summary of Prior work

Prior work in this area includes algorithms applied to other ICU classifier events such as unplanned intubation in Trauma ICU patients (9), moving of patients to the ICU within 12 hours (10,11), as well as the likelihood of, mortality due to, or survival despite events similar to VACs such as sepsis (12–15), acute respiratory distress syndrome (ARDS), and respiratory failure after initiation of extracorporeal membrane oxygenation (ECMO) treatment (9). Such studies have produced results with a wide range of AUCs ranging between 0.625 and 0.92. A study by Huber et al., 2020 (16) compared various ARDS definitions and associated information during intubation and had a range of AUC scores from 0.620 to 0.824 depending on features used for prediction. Though studies predicting VACs are few, there has been work over related predictive analyses. For example, the APACHE II score has been used to predict mortality from VAP with an AUC of .81 (9) and VAE definition conditions have been shown to predict poorer outcomes in sepsis patients using a log-rank test (15).

Most hospital admission centers calculate critical patient scores within the first 24 hours of ICU admission (10). There is a clear need for a prediction model regarding events prior to patient decompensation, specific to mechanical ventilation (16). Other predictive models, such as the likelihood of sepsis, use a similar approach (12–15). When prognosticating, most clinicians are more concerned about giving positive prognosis (and altering treatment accordingly) for patients who would otherwise get a complication, favoring a model with higher sensitivity. Prior research indicates both a need for and the potential to create a predictive model that meets these specifications.

### Hypotheses

For both cohorts, we built random forest models using manual as well as tsfresh low-frequency features. For the high frequency cohort, we also built a random forest model using the top tsfresh high-frequency features.

The main variables we were interested in comparing were cohort size, temporal resolution of data, and automated features. In examining model performance while varying these different aspects, we hypothesized that:

1. Increasing temporal resolution through use of waveform data will improve a random forest model’s ability to predict VACs
2. Increasing the sample size of the training set will improve a random forest model’s ability to predict VACs
3. Use of tsfresh in generating features will improve a random forest model’s ability to predict VACs relative to a model with manually-derived features

## METHODS

### Study Populations

We selected patients retrospectively from the MIMIC-III dataset. Published by the Massachusetts Institute of Technology (MIT) and Physionet, MIMIC-III contains 46,338 adult and pediatric patients admitted to the critical care units of Beth Isreal Deaconess Medical Center in Boston, Massachusetts between 2001 and 2012. Adult patients were included if they received mechanical ventilation for at least 96 hours – the minimum time to be formally at risk for a VAC. Only one ICU stay as considered per patient. Patients with more than 40% of missing low-frequency data were excluded (Figure 1). Patients with sufficient data for statistical feature generation were placed in Cohort One. Patients with sufficient physiologic time series data were placed in Cohort Two.

**Figure 1:**
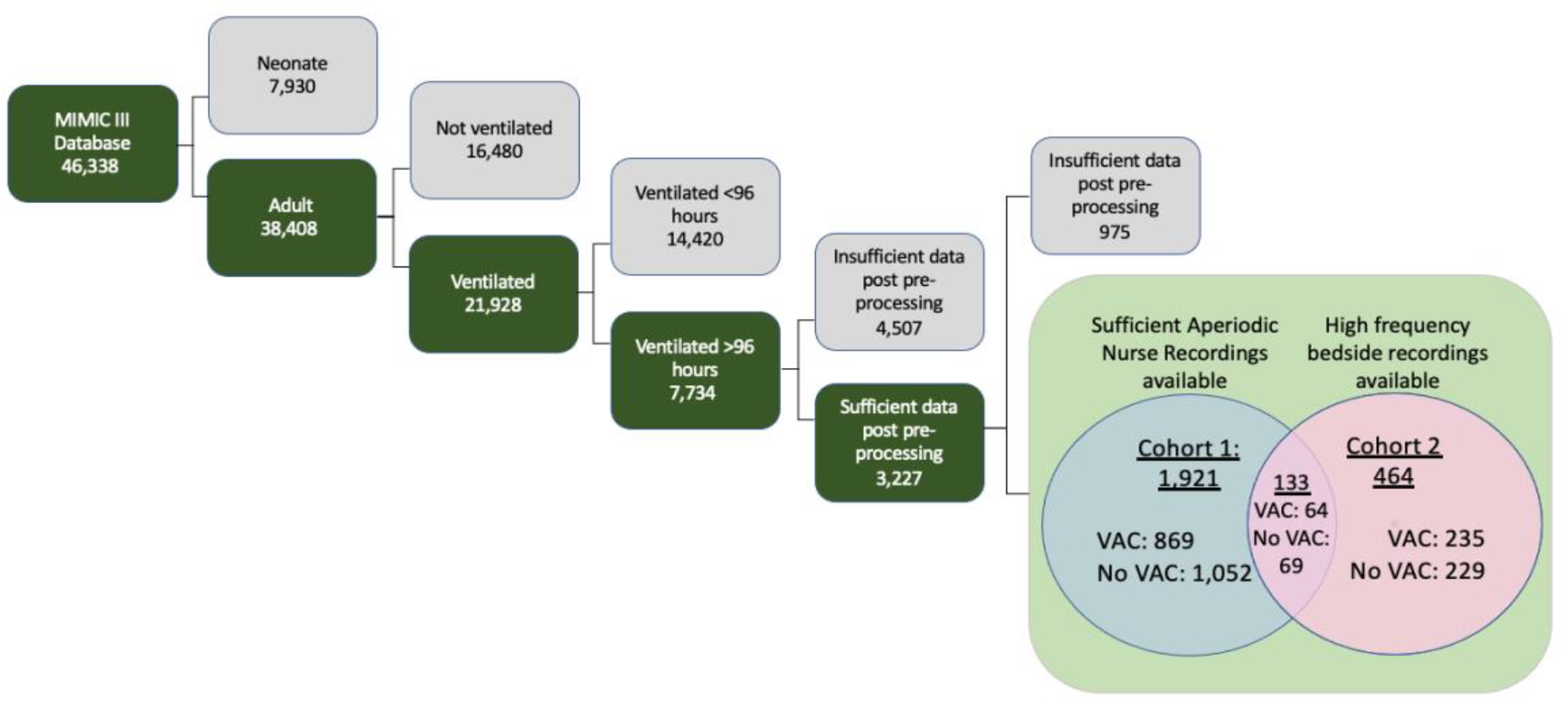
Inclusion criteria for MIMIC-III database to arrive at anticipated patient Cohorts One and Two, both of which were composed of patients who had enough data points for automatic statistical feature extraction methods. Cohort One consists of patients with aperiodic data recorded by nurses and clinicians, and Cohort Two consists of patients with high frequency waveform data.

### Prediction Task

We considered the outcome of VAC, as defined by the CDC to be 48 hours of stable or decreasing daily minimum FiO2 and PEEP settings, followed by 48 hours where either the daily minimum FiO2 or PEEP increased by 0.2 or 3 mm Hg respectively. The time of transition from stable/decreasing settings to increasing settings was considered the VAC onset time. For patients at risk of a VAC (having been ventilated for at least 48 hours), we then considered the binary prediction at hour of mechanical ventilation with hours labeled as 1 for patients in which a VAC onset occurred and 0 otherwise. Models considered patient demographic variables and patient data from the previous 35 hours (chosen a priori) to one hour before VAC onset to make a prediction.

### Data Preprocessing

For each patient, demographics (age, gender), labs (arterial pH, PaO_2_, glucose, WBC), nurse recorded vital signs (temperature, heart rate, blood pressure, respiratory rate, SpO2), nurse recorded ventilator settings/measurements (PEEP, mean airway pressure), and physiologic time series data were extracted. Physiologically implausible values were removed (Table 2).

Low-frequency values missing for more than one hour were replaced with the mean value from the training set. High-frequency data were partitioned into five minute intervals for median down-sampling. After down-sampling, data gaps longer than one hour (12 consecutive missing values) were filled by linear interpolation in a manner that ensured causality. Data gaps shorter than 1 hour were carried forward from the most recent non-null value. For leading null values in the window that could not be replaced using carry-forward (all prior values before the window were also null), the value was backward-filled with the first future non-missing value.

### Feature Extraction

Different time frames of data before VAC onset were tested for model input with preliminary models, with the best results found using the data 36 hours before VAC onset. Data was therefore extracted 36 hours before VAC onset, and features were extracted during the first 35 hours of this data, allowing for prediction one hour before VAC onset. Variables with 40% or more missing samples across patients were removed. We computed statistical summaries (mean, variance, number of features, min, max, and range) of 12 hours windows and of three hour windows of all variables, resulting in 278 features. We also used the tsfresh package (16) to automatically generate features, resulting in 916 features from 20 low frequency signals and 12853 features from 6 high frequency signals.

### Feature Selection

Highly correlated features were removed when Pearson correlations were greater than 0.95. Forward selection, a stepwise regression technique utilizing significance (p-value) levels from model performance, was used to select the most relevant features from each type of data, resulting in a top 35 tsfresh features and 29 manual features from the low frequency data, as well as a top 167 tsfresh features from the high frequency data.

### Model Training and Validation

We trained and compared five different models using random forest classification, as implemented by the scikit-learn library (17) in python 3. For Cohort One, two models were created: one using tsfresh features derived from low-frequency nurse recorded data, and one using manually defined features from the same. For Cohort Two with high frequency data availability, three models were created: one using tsfresh features derived from low-frequency data, one using tsfresh features derived from high-frequency data, and one using manual features from low-frequency data.

The models constructed are summarized below:

1. Tsfresh features derived from low frequency data of 1921 Cohort One patients
2. Tsfresh features derived from low frequency data of 464 Cohort Two patients
3. Tsfresh features derived from high frequency data of 464 Cohort Two patients
4. Manual features derived from low frequency data of 1921 Cohort One patients
5. Manual features derived from low frequency data of 464 Cohort Two patients

Random forests have 4 hyperparameters – the number of trees, the maximum features, the maximum tree depth, and splitting criterion. A hyperparameter search was conducted with the GridSearchCV function from scikit-learn. Model performance was evaluated using 10-fold cross validation (e.g. for hyperparameter setting 10 different models were built using 90% of the data and performance was evaluated on the remaining 10% of the data, then averaged). Model performance was measured by the AUROC. The best performing hyperparameters were then reported as is.

Random forest, our choice of classification, is a machine learning algorithm that classifies testing data based on uncorrelated decision trees (set at 512 trees), each of which intuitively ask a sequence of questions about the data until it arrives at a classification. A hyperparameter search gave optimal parameters of ‘auto’ for max features, 500 estimators, a max depth of 9, and the Gini criterion. However, these settings did not cause model performance to materially deviate from default settings.

Random forest models were constructed using features chosen after feature reduction and selective feature elimination. Model outcome was incidence of a VAC during the patient’s ICU stay. Classification results were obtained from K-folds cross-validation (k = 10), and summary statistics from the average area under the receiver operating characteristic (AUROC) curves are reported in Table 3.

## RESULTS

Cohort One, patients with sufficient EHR data for statistical feature generation, included 1921 patients experiencing 869 VACs. Cohort Two, patients with sufficient physiologic time series data for statistical feature generation, included 464 patient experiencing 235 VACs. (133 patients were in both cohorts). Table 1 describes the characteristics of these cohorts.

**Table 1:** VAC Cohort Patients.

|  | Cohort 1 | Cohort 2 |
| --- | --- | --- |
| *Unique Subjects (n)* | 1921 | 464 |
| *Age, years (mean, (s.d.))* | 69.3, (43.49%) | 66.3, (36.6%) |
| *Male gender (n, (%))* | 1126, (58.7%) | 272, (58.6%) |
| *Ventilator Duration (mean hours)* | 236 | 206 |
| *Length of ICU stay (mean days)* | 13.84 | 12.03 |
| *Hospital mortality (n, (%))* | 637, (33.2%) | 154, (33.2%) |
| *Patients with VAC* | 869 | 235 |

### Feature Generation and Selection

Tsfresh generated 916 statistical features from EHR data in cohort one. 350 features with Pearson correlation greater than 0.95 were removed. 531 additional features were removed by forward selection, resulting in 35 tsfresh features on low-resolution physiologic data. On cohort two, tsfresh generated 12,853 statistical features from PTS data, of which 12,491 were removed by Pearson correlation and 195 were removed by forward selection, resulting in 167 tsfresh features on high-resolution physiologic data.

### Model Performance

Table 3 and Figure 2 summarize model performance (AUROC and accuracy) of the five models using 10-fold cross validation. On Cohort One, automated low-resolution features (M1) achieved an AUC of 0.762 ± 0.002 and manual features (M4) achieved an AUROC of 0.792 ± 0.001. On cohort two, automated high-resolution features (M3) achieved an AUROC of 0.826 ± 0.006, automated low-resolution features (M2) achieved an AUROC of 0.626 ± 0.007, and manual features (M5) achieved an AUROC of 0.737 ± 0.002.

**Table 2:** Valid physiological vital signs parameter ranges to determine outliers. Vital signals include heart rate (HR), respiratory rate (RR), diastolic and systolic blood pressure (B.P.), oxygen saturation (SpO2), positive end-expiratory pressure (PEEP), and fraction of inspired oxygen (FiO2).

|  | HR<br>(bpm) | RR<br>(breaths/min) | Diastolic<br>B.P.<br>(mmHg) | Systolic<br>B.P.<br>(mmHg) | SpO2<br>(%) | PEEP<br>(cmH2O) | FiO2<br>(%) |
| --- | --- | --- | --- | --- | --- | --- | --- |
| *Lower bound* | 30 | 6 | 20 | 50 | 80 | 0 | 21 |
| *Upper bound* | 200 | 50 | 120 | 220 | 100 | 20 | 100 |

**Table 3:** Summary of Model Results. Number of patients indicates the cohort used for both training and testing of models. The number of features column represents the number of features found to give the largest AUC, and was found using the forward selection technique for feature selection.

| Model | Number of<br>patients | Number of<br>features | AUC<br>mean | AUC<br>variance | Accuracy<br>mean | Accuracy<br>variance |
| --- | --- | --- | --- | --- | --- | --- |
| ***M1** EVENTS*<br>*tsfresh features* | 1921<br>(Cohort 1) | 35 | .7620 | .0017 | .6976 | .0012 |
| ***M2** EVENTS*<br>*tsfresh features* | 464<br>(Cohort 2) | 35 | .6262 | .0069 | .5901 | .0066 |
| ***M3** PTS data*<br>*tsfresh features* | 464<br>(Cohort 2) | 167 | .8257 | .0055 | .7650 | .0075 |
| ***M4** Manual*<br>*features* | 1921<br>(Cohort 1) | 29 | .7916 | .0010 | .7314 | .0011 |
| ***M5** Manual*<br>*features* | 464<br>(Cohort 2) | 29 | .7373 | .0023 | .6811 | .0019 |

**Figure 2:**
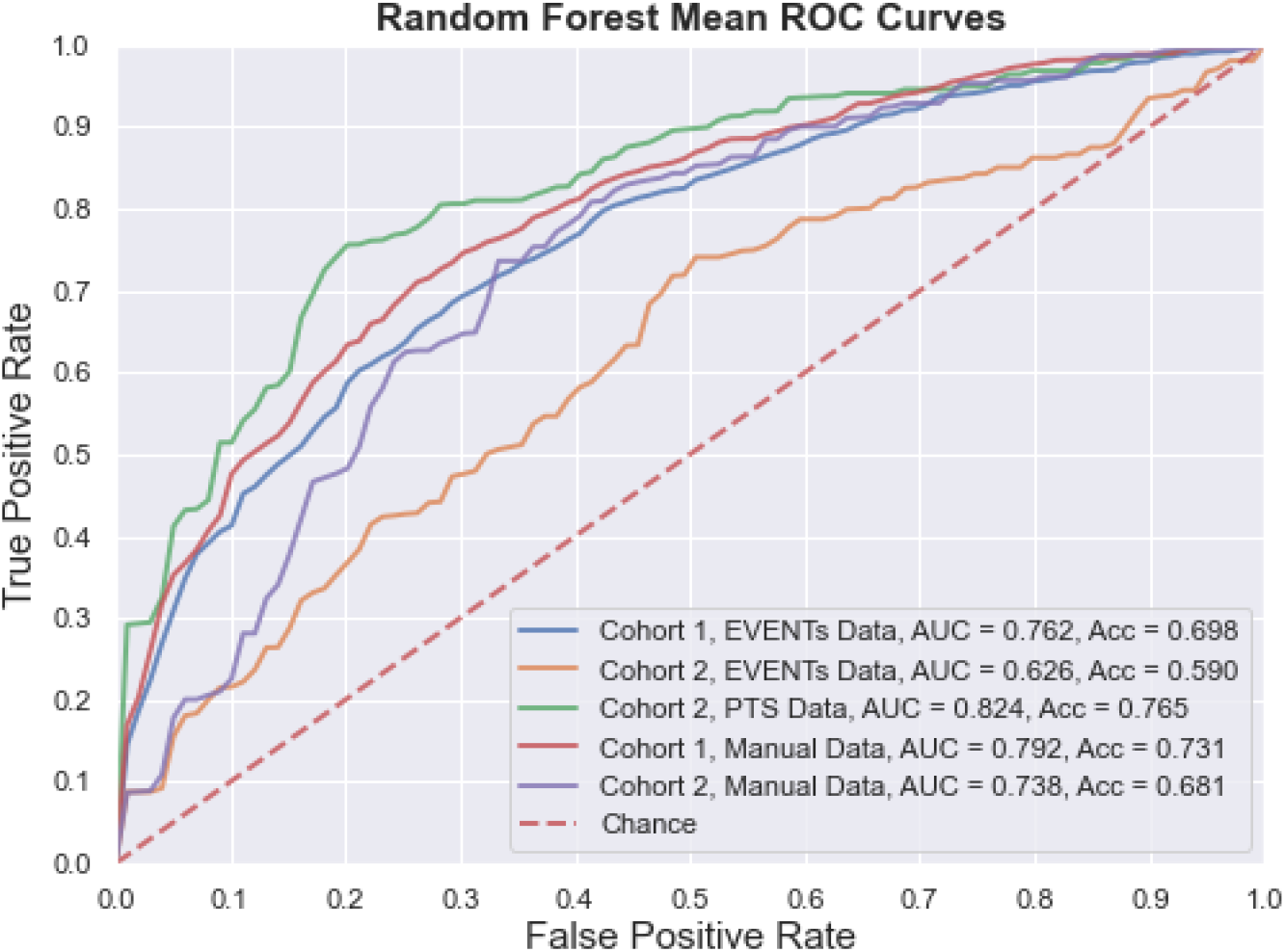
Plot of Random Forest Mean AUCs

**Figure 3:**
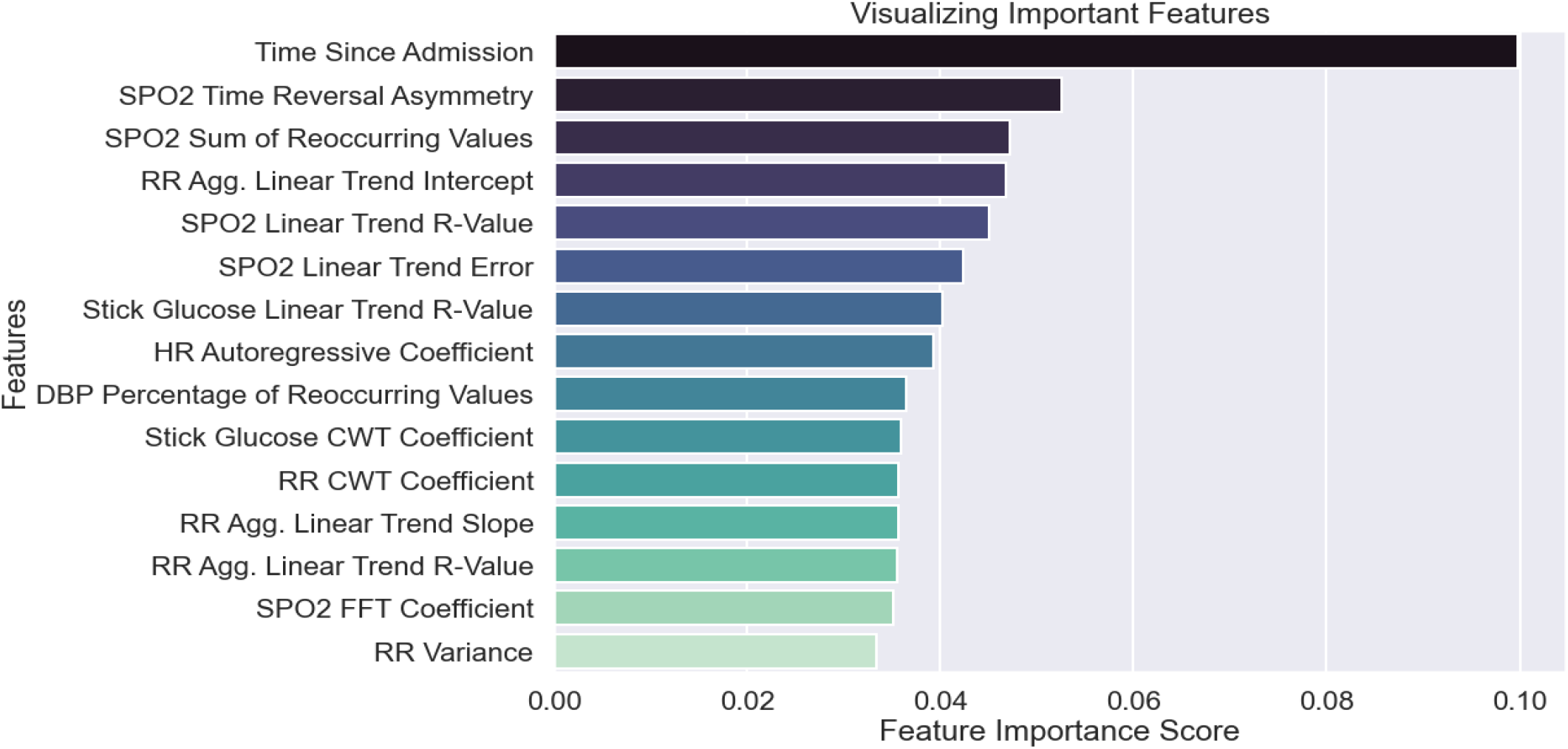
Top 15 significant Features from Cohort 1 tsfresh EVENTs features (M1)

**Figure 4:**
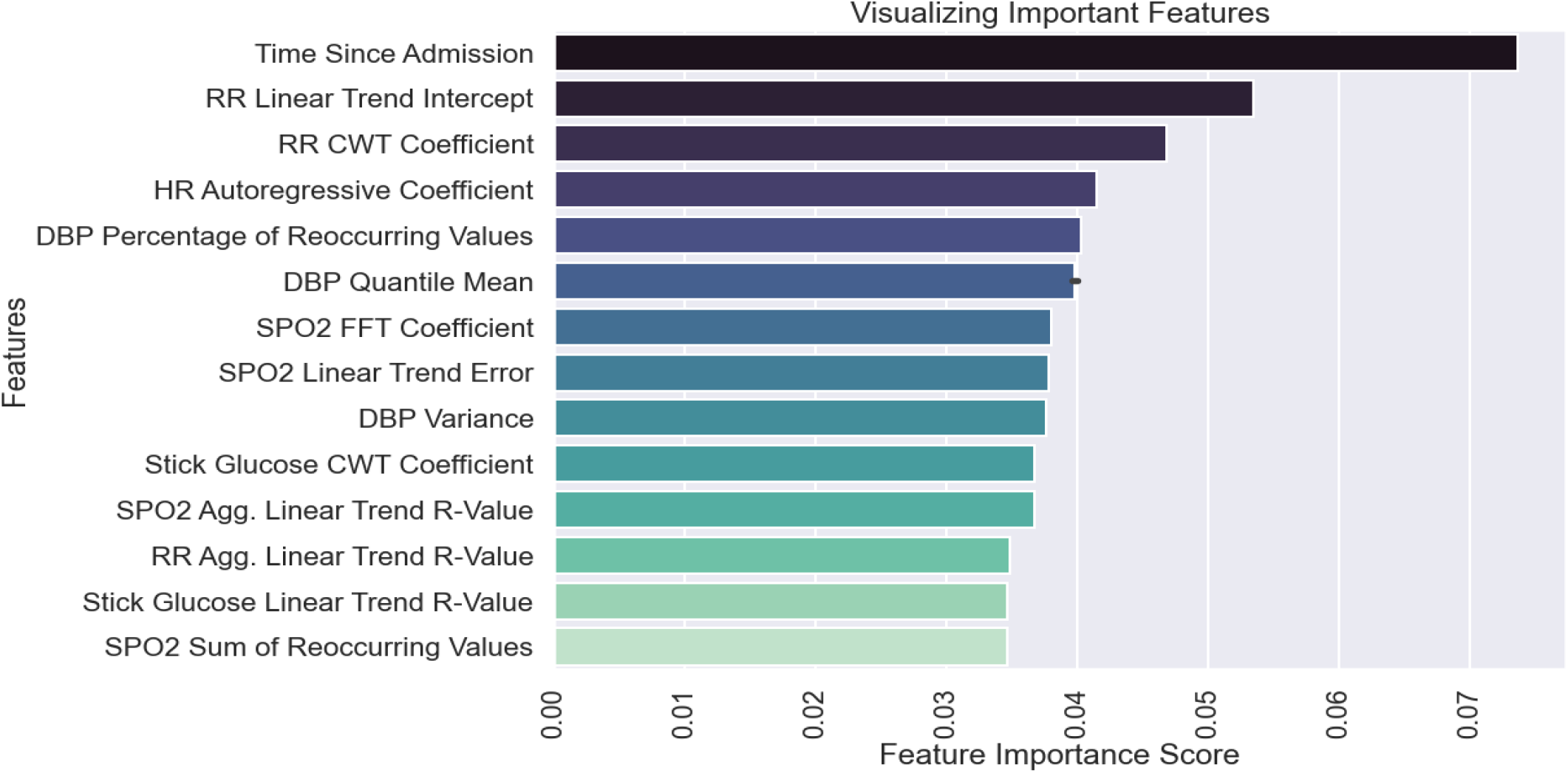
Top 15 significant Features from Cohort 2 tsfresh EVENTs features (M2)

**Figure 5:**
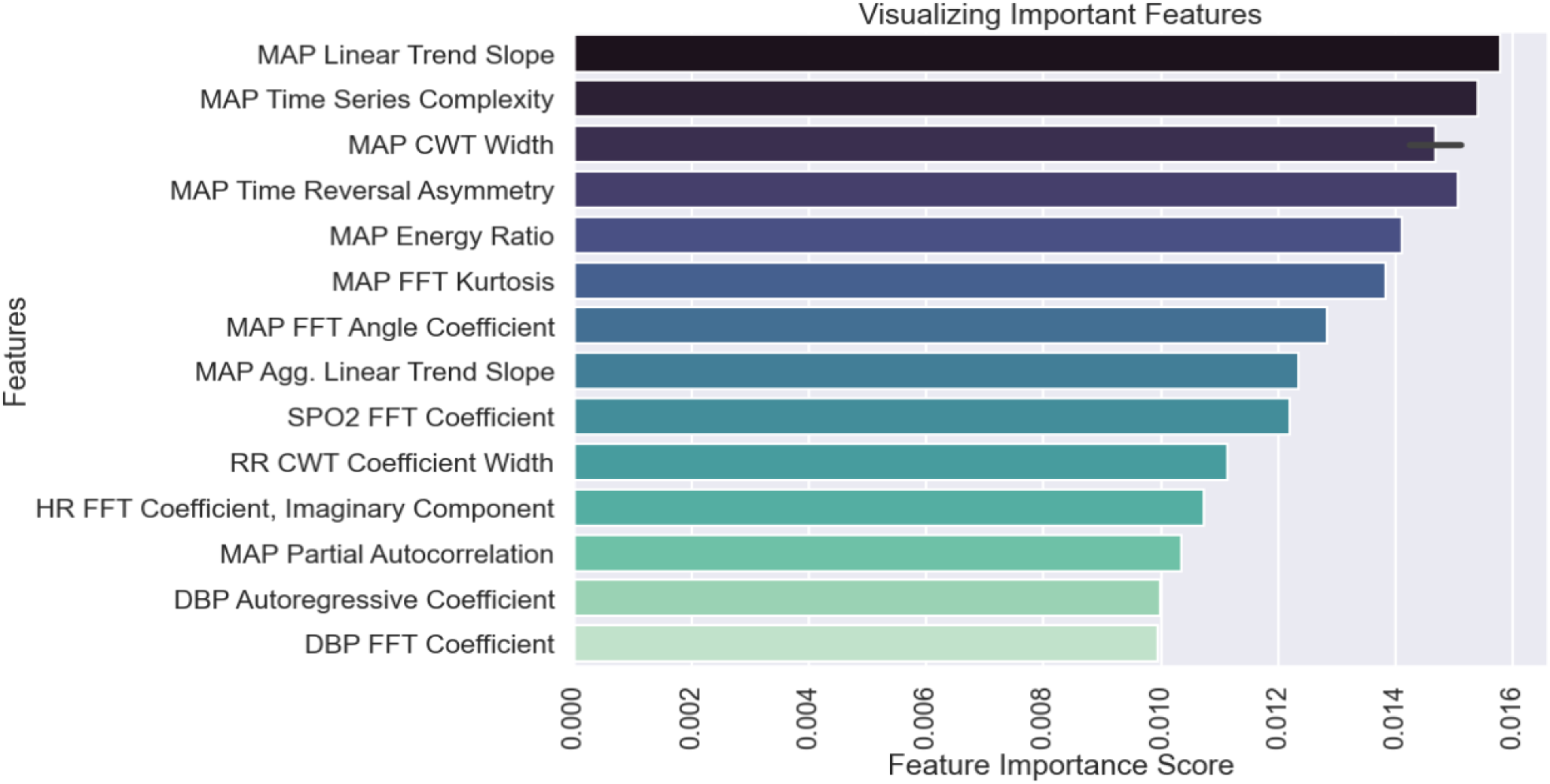
Top 15 significant features from PTS data (M3)

**Figure 6:**
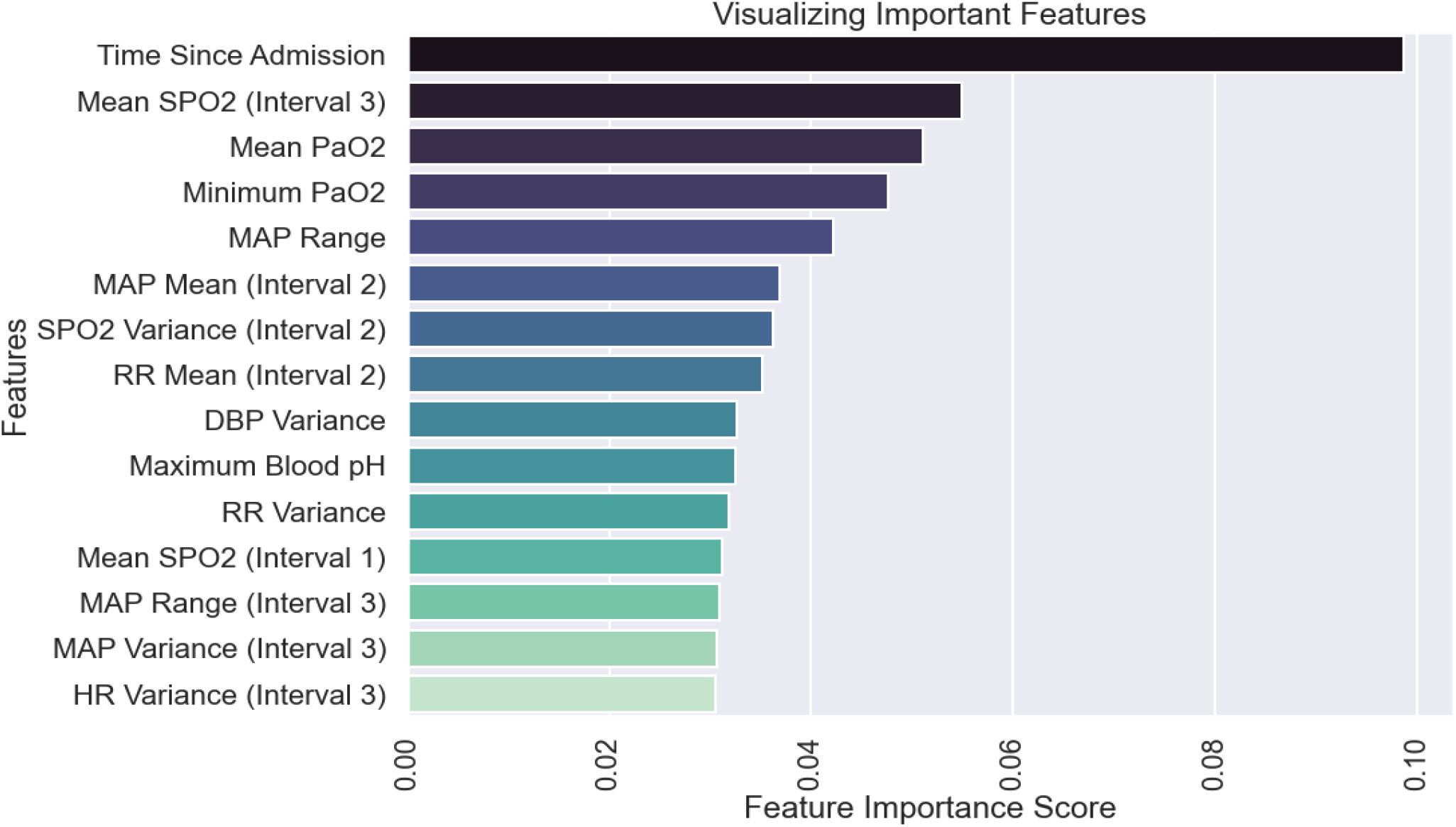
Top 15 significant Features from Cohort 1 manually derived features (M4)

**Figure 7:**
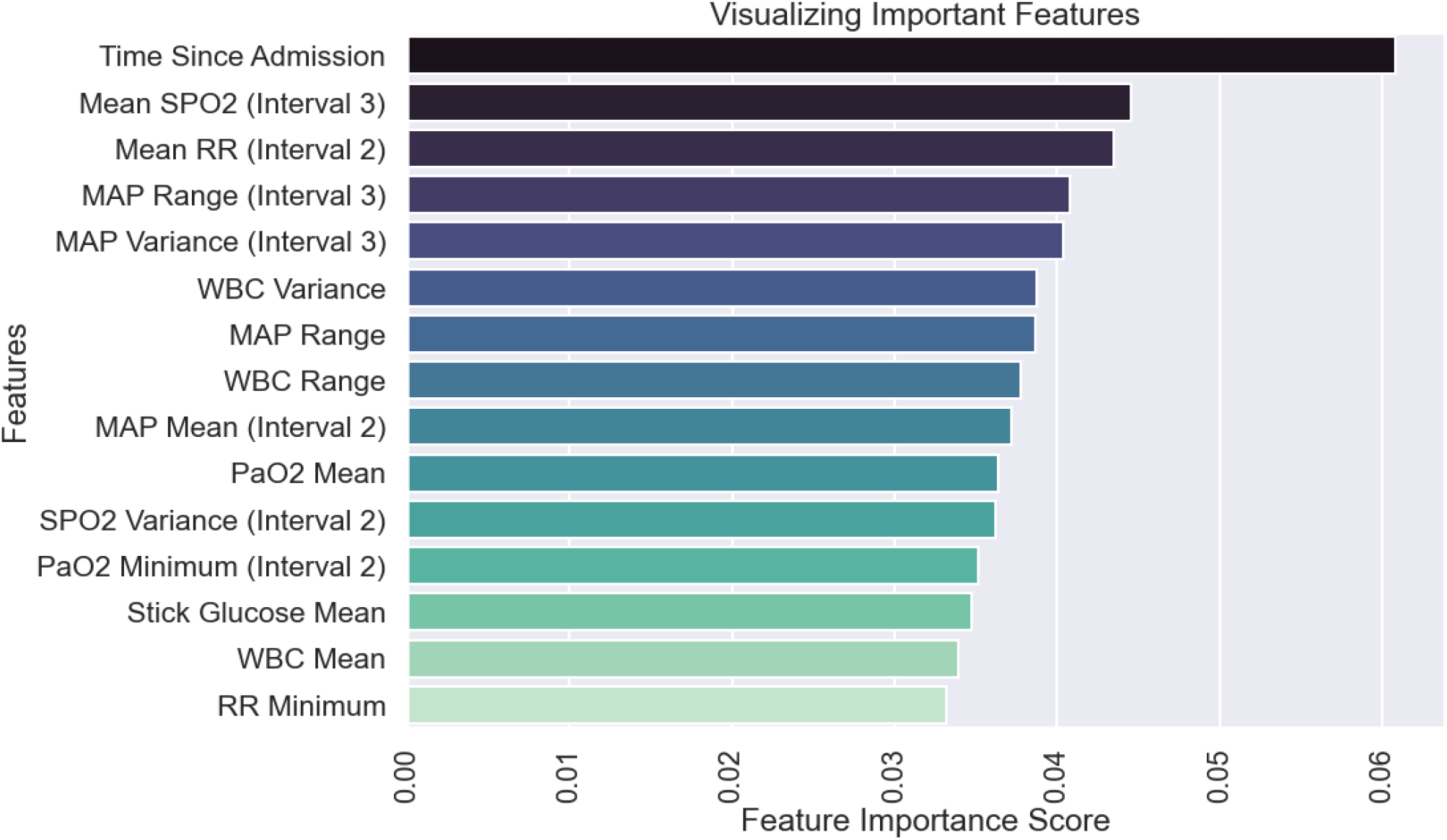
Top 15 significant Features from Cohort 2 manually derived features (M5)

### Feature Importance

These most relevant features constitute a set of possible baseline risk factors, and derive from the signals of airway pressure, SPO_2_, respiratory rate (RR), diastolic blood pressure (DBP), systolic blood pressure (SBP), heart rate (HR), PaO_2_, glucose stick levels, mean arterial pressure (MAP), Glasgow coma scale score (GSC score), temperature (TEMP), and white blood cell count (WBC). Additional static features included a binary categorical variable for if a neuromuscular blocker was prescribed, shock index (HR/SBP), age, gender, time since admission to the ICU, and the number of vital sign measurements recorded in a patient’s data collection window.

## DISCUSSION

The CDC ventilator associated complication (VAC) surveillance criteria identifies a range of complications associated with mechanical ventilation in adult patients (18) and has been associated with increased morbidity/mortality (6). Applying machine learning to patient demographic data and routinely measured physiologic time series data, we demonstrate VACs can be predicted with an AUROC of 0.826 one hour before they occur (and 49 hours before the CDC surveillance definition can be evaluated).

Clinically, model outputs can be applied in at least three ways. First, when an individual patient’s score crosses a threshold associated with high specificity, immediate diagnostics and interventions could be triggered to assess for early signs of complications (such as pneumothorax) and optimize pulmonary function (such as more aggressive pulmonary toilet). Second, an individual patient’s score trajectory over time may provide early warning about occult developments that may progress into decompensation. Finally, relative scores in a cohort of patients, such as ICU provider’s panel, could help triage both attention and resources to patients at greatest risk of decompensation.

Methodologically, we demonstrate the viability of automated feature generation from high-resolution physiologic time series data. Packages like tsfresh can generate a large number of non-linear summarizations of physiologic time series data, some more related to the outcome than manually constructed features. After removal of (a) features highly correlated with other features and (b) features poorly correlated with the outcome while controlling for other features, model performance (AUROC 0.826 for M3) exceeds that of manual features (AUROC 0.737 for M5). This result did not hold for automated features generated from low resolution data, where performance was consistently lower than for models built with manual features (M1 vs M4 and M2 vs M5) and lower than for models built with automated features from high-resolution. We also demonstrate, not unexpectedly, that models with identical features but trained on more patients have better performance (M1 vs M2 and M4 vs M5).

The feature importance provides insight into the models. Time since admission was the most important feature in all models, except for M3 which used automated features from the high-frequency PTS data. While random forest variable importance does not indicate the variable’s relationship with the outcome, this is consistent with published data that VAC risk increases over time (4). Model M3’s ability to provide better predictions without relying on time since admission indicates it has found physiologic features more closely associated with VAC than time since admission. The top eight features of model M3 are derived from MAP, followed by signals derived from HR, RR, DBP, and SPO_2._ In the best performing manual model, M4, mean SpO2 in last interval is 2^nd^ most important, followed by PaO2 statistics, then MAP statistics.

Strengths of our approach include the use of VAC as a prediction target. VAC has been demonstrated to be a clinically valid construct for patient decompensation while ventilated that is associated with worse outcomes. The feature generation and selection methodology is scalable to dense time series data. Compared to manual feature engineering, it is able to find more informative features that improve model outcome. While the automated features are not as widely recognized as standard statistical features, they are defined transformations that can be analyzed. For example, FFT and CWT coefficients correspond to specific frequencies or patterns occurring in the data. This analyzability stands in contrast to deep learning approaches where learned features are inherent to the neural network structure and cannot be easily analyzed. Finally, our use of the MIMIC cohort makes this analysis easy to reproduce and compare with other approaches.

Limitations include that the patient cohort is from a single institution, the small number of patients with sufficient data prevented evaluation of the developed algorithm on a held-out dataset, and the lack of an external validation. Additionally, the variables that define a VAC, while more objective than those used for the previous VAP construct (2) are under clinician control and rely on timely adjustment. Failure to wean FiO2/PEEP while a patient is improving could mask a subsequent VAC. Similarly, escalation of FiO2 and PEEP off the ARDSnet protocol (19) could trigger a VAC. One solution would be to balance these settings with a measure of oxygenation, such as PaO2 or SpO2. Finally, the VAC focuses primarily on oxygenation, and may miss decompensation that requires increased ventilation.

This work raises several questions for future work. First, given that automated feature generation outperformed manual features on the high frequency data, where does the frequency cut-off occur, and can higher frequencies further improve performance. Second, can automated features be made more interpretable.

### Future Directions

This model is still capable of improvement. Application of neural networks to raw waveform data rather than uniform down-sampling of the physiological time series data may provide greater accuracy or earlier predictions for real-time usage in the ICU. This model can also be externally validated on other critical care databases as to determine its functionality across different datasets. With such improvements, a model could be created to provide an hourly risk score of the patient’s status, or a sliding time window for clinicians to understand when a patient’s vital signs worsen, as to signal the need for appropriate clinical interventions.

## CONCLUSION

We developed risk prediction models specific to the mechanically ventilated ICU population. The models’ ability to classify patients accurately compares favorably to the current classification standard using APACHE risk scores (10). Though this model relies on existing records, the methodology suggests feasibility in further use of high-frequency data. Future work using more computationally intensive models and higher-frequency patient data may increase relevant classification metrics even further.

## Data Availability

All data produced in the present work are available upon reasonable request to the authors.

